# International stakeholder engagement on clinical care and research for nocturnal hypoxaemia in pulmonary fibrosis

**DOI:** 10.1101/2025.04.14.25325780

**Authors:** Leona Dowman, Shane Landry, Natasha Smallwood, Catharina Moor, Bradley Edwards, Christopher J Ryerson, Christine F McDonald, Nicole Goh, Harry Patsamanis, Lynne Cochrane, Simon Joosten, Marlies Wijsenbeek, Magnus Ekström, Sebastian V Moreno, Graham Hepworth, Anne E Holland, Yet H Khor

## Abstract

**Background:** Nocturnal hypoxaemia is common in people with pulmonary fibrosis (PF). This study aimed to explore current practice and establish global perspective on research priorities and outcome measures of importance in sleep assessment and treatment of nocturnal hypoxaemia for PF.

**Methods:** People with PF, as well as healthcare professionals (HCPs) including clinicians and researchers with expertise in PF, sleep medicine, and oxygen therapy, were recruited internationally to participate in a mixed-methods online survey followed by online focus groups.

**Results:** A total of 68 people with PF and 73 HCPs from 29 countries completed the survey, with 14 and 36 joining the focus groups, respectively. 51% of patients had previous sleep assessment, with 75% of the remaining expressing its need as part of the disease assessment for PF. 64% of HCPs performed sleep assessments routinely or as clinically indicated, with 82% indicating the assessment being very or somewhat important. The top research priority from people with PF and HCPs was treatment effects of nocturnal hypoxaemia on symptom burden (including health-related quality of life [HRQoL]). Other key research priorities identified were safety and tolerability of nocturnal oxygen therapy, treatment effects of nocturnal hypoxaemia on mortality and pulmonary hypertension, diagnostic approaches for assessing sleep and nocturnal hypoxaemia, predictors of nocturnal hypoxaemia, developing a user-friendly oxygen therapy device, and patient awareness of the significance of nocturnal hypoxaemia. For outcome measures of importance, both groups prioritised HRQoL. In addition, people with PF highly ranked forced vital capacity, nocturnal oxygenation status, and apnoea-hypopnoea index, while HCPs selected long-term sequelae such as survival and development of PH. Impact of nocturnal hypoxaemia and sleep disturbance on cognitive performance was raised by people with PF as a key research topic, which was agreed by HCPs.

**Conclusion:** This study provides important insights into stakeholders’ priorities to guide future research on sleep assessment and treatment of nocturnal hypoxaemia in PF.

## INTRODUCTION

Nocturnal hypoxaemia is a significant clinical manifestation of pulmonary fibrosis (PF), also termed as fibrotic interstitial lung disease, affecting approximately a third of patients.^1,2^ People with PF are susceptible to developing nocturnal hypoxaemia with reduced ventilation and impaired gas exchange that worsen during sleep.^3^ In addition, obstructive sleep apnoea (OSA) is a common comorbidity in people with pulmonary fibrosis,^3^ which results in intermittent hypoxia during sleep due to repetitive pharyngeal narrowing and collapse. Nocturnal hypoxaemia is associated with worsened fatigue, daytime functioning, health-related quality of life (HRQoL), and survival, as well as the development and presence of pulmonary hypertension (PH) in people with pulmonary fibrosis.^1,2,4^

Treating nocturnal hypoxaemia has the potential to improve health outcomes in people with f pulmonary fibrosis. Nocturnal oxygen therapy (NOT) is a therapeutic option, however, no randomised controlled trials have evaluated its efficacy in this population. As a result, there is wide variability in the recommended diagnostic and prescribing criteria for NOT in PF across international guidelines.^5,6^ Furthermore, there is a lack of guidance on sleep assessment in people with pulmonary fibrosis, which is necessary for the diagnosis of nocturnal hypoxaemia. To advance understanding of clinical care and inform future research for nocturnal hypoxaemia in pulmonary fibrosis, this international study of patients and healthcare professionals (HCPs) aimed to explore current perceptions and practice on sleep assessment and prescription of NOT, to identify key research questions on nocturnal hypoxaemia and NOT, and to establish key outcome measures for future research of NOT.

## METHODS

### Study design

This international study consisted of mixed-methods surveys, which was followed by focus groups for validating findings. Ethics approval was received from Monash University Human Research Ethics Committee (Project ID: 44114).

### Study participants

There were two groups of participants: (1) people with pulmonary fibrosis, and (2) HCPs, including clinician and researchers, with expertise in pulmonary fibrosis, sleep medicine and/or oxygen therapy. People with PF were recruited using an electronic flyer circulated by the Lung Foundation Australia, European Pulmonary Fibrosis Federation, and PF Warriors. For HCPs, a purposive sample was identified through clinical practice guidelines, peer-reviewed literature, and professional networks, with the aim to achieve diverse representation of age, sex, career stage, profession backgrounds, and geographical practice locations. People with PF and HCPs who completed the surveys were invited to participate in the online focus groups.

### Mixed-methods surveys

The online mixed-methods surveys were purpose-built using Qualtrics and distributed between September and November 2024, with separate questions being designed for people with PF (**Online Supplementary: Appendix 1**) and the HCPs (**Online Supplementary: Appendix 2**). Each survey consisted of three sections related to: 1) experiences and views regarding sleep assessment and treatment for nocturnal hypoxaemia in pulmonary fibrosis, 2) top research priorities on nocturnal hypoxaemia in PF (up to three nominations), and 3) a list of outcome measures for assessing treatment effects of nocturnal oxygen therapy in pulmonary fibrosis. Items included in the evaluation of outcome measures for nocturnal oxygen therapy were identified from a systematic review and investigators’ input,^1^ with participants being invited to nominate additional relevant outcomes that were not listed.

The survey contained both quantitative questions with multiple choice responses and a Likert scale, and qualitative questions with free text responses, as appropriate. The Likert scale used was 5 points, with 0 “not important at all”, 1 “somewhat unimportant”, 2 “neutral”, 3 “somewhat important”, and 4 “very important”. Participant demographics collected were age and sex for both participant groups, with additional information on country of residence, disease type and years of diagnosis for people with PF and professional role, area of expertise, country and years of practice for HCPs.

### Online focus groups

After completing analyses for surveys, online focus groups were conducted in December 2024 using Zoom teleconference software, which were facilitated by investigators. The survey results for both people with PF and HCPs were presented, and focus group participants were asked to review and verify findings on key research questions on nocturnal hypoxaemia and outcome measures for evaluation of nocturnal oxygen therapy in pulmonary fibrosis. Open-ended questions and discussions were encouraged. Sessions were audio recorded and transcribed verbatim. Similar to the surveys, participant demographic information was collected to establish their representation.

### Data analysis

For the mixed-methods surveys, quantitative questions were summarised using counts and frequencies for multiple choice responses, and median with interquartile range and percentages for “very important” for Likert scale responses. The free text responses were analysed using a content analysis approach by two investigators,^7^ who independently coded the responses and subsequently grouped through iterative discussion. Transcripts from the online focus groups were similarly coded by two investigators independently and then agreed upon by consensus.

## RESULTS

A total of 68 people with PF and 73 HCPs completed the mixed-methods surveys (**Tables 1 and 2**). There was diverse representation of people with PF and HCPs from across the world, with greatest participation from Australia and Europe, respectively. The complete breakdown by country is shown in **Figure 1**. Both participant groups had equal gender distribution. People with PF were spread across a range of age categories, with 40% being diagnosed >5 years ago and 25% reporting current or previous use of nocturnal oxygen therapy. Most HCPs were physicians (88%) and/or researchers (30%). The vast majority of HPCs specialised in PF (90%), followed by oxygen therapy (27%) and sleep medicine (16%). 68% of HCPs had over 10 years of working experience. There were 13 people with PF and 30 HCPs who participated in the online focus groups, with similar characteristics compared to the overall groups (**Tables 1 and 2**).

**Table 1.**
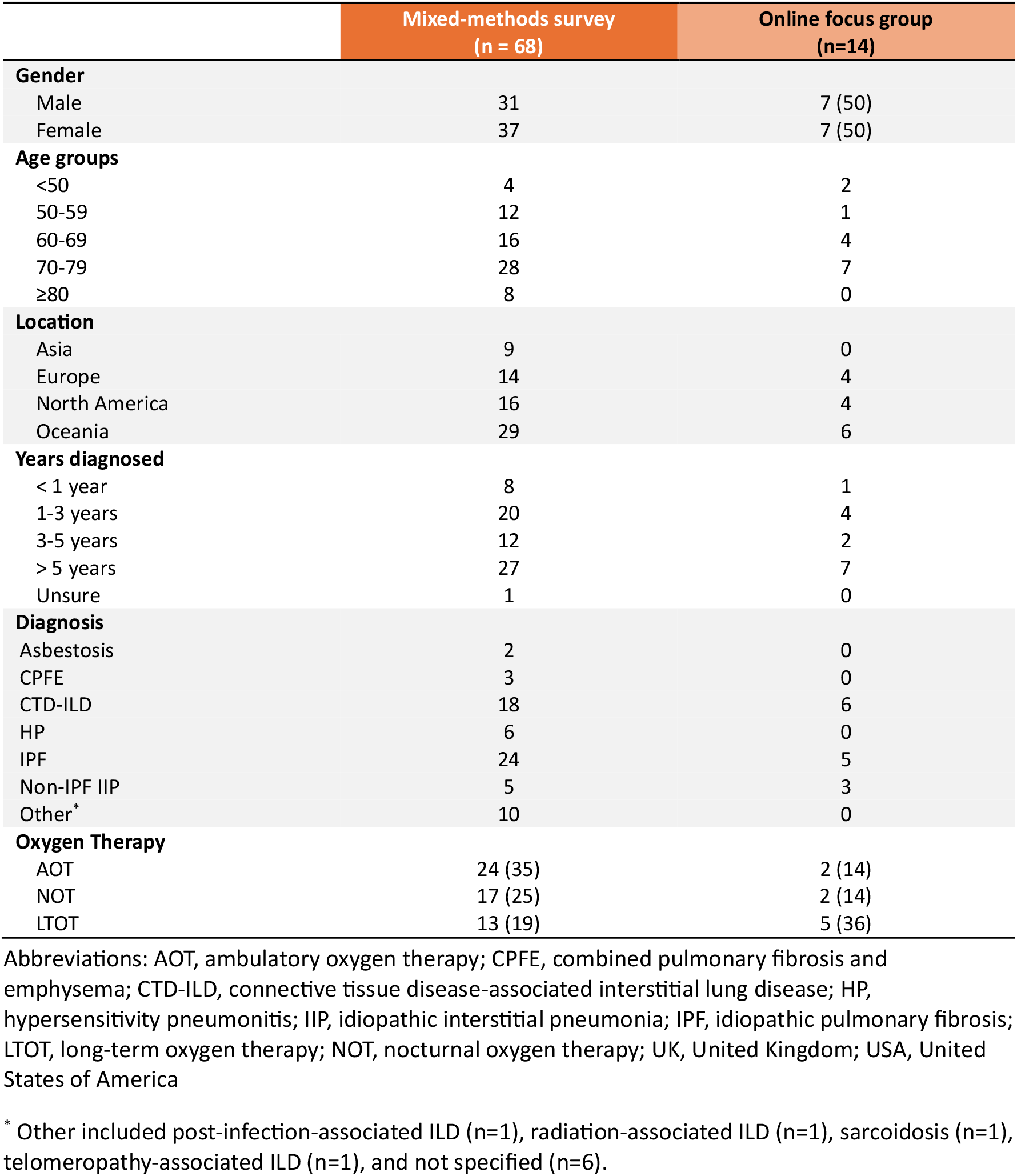
Characteristics of people with pulmonary fibrosis.

**Table 2.** Characteristics of healthcare professionals.

|  | Mixed-methods survey<br>(n = 73) | Online focus group<br>(n=39) |
| --- | --- | --- |
| **Gender** |  |  |
| Male | 36 (49) | 21 |
| Female | 37 (51) | 18 |
| **Age groups (years)** |  |  |
| 26-35 | 6 (8) | 2 |
| 36-45 | 31 (42) | 15 |
| 46-55 | 20 (27) | 12 |
| > 55 | 16 (22) | 10 |
| **Location** |  |  |
| Asia | 16 | 13 |
| Europe | 26 | 9 |
| North America | 20 | 11 |
| Oceania | 10 | 5 |
| South America | 1 | 1 |
| **Occupation\*** |  |  |
| Allied health professional | 1 | 0 (0) |
| Nurse | 6 | 3 (8) |
| Physician | 64 | 36 (92) |
| Researcher | 23 | 20 (51) |
| **Area of expertise\*** |  |  |
| ILD | 66 | 35 (90) |
| Oxygen therapy | 20 | 11 (28) |
| Sleep medicine | 13 | 9 (23) |
| **Clinical/research experience (years)** |  |  |
| ≤ 5 | 3 | 2 |
| 6-10 | 19 | 6 |
| 11-20 | 27 | 16 |
| 21-30 | 15 | 9 |
| > 30 | 9 | 6 |

Abbreviations: ILD, interstitial lung disease; UK, United Kingdom; USA, United States of America
\* Respondents could select more than 1 category

**Figure 1.**
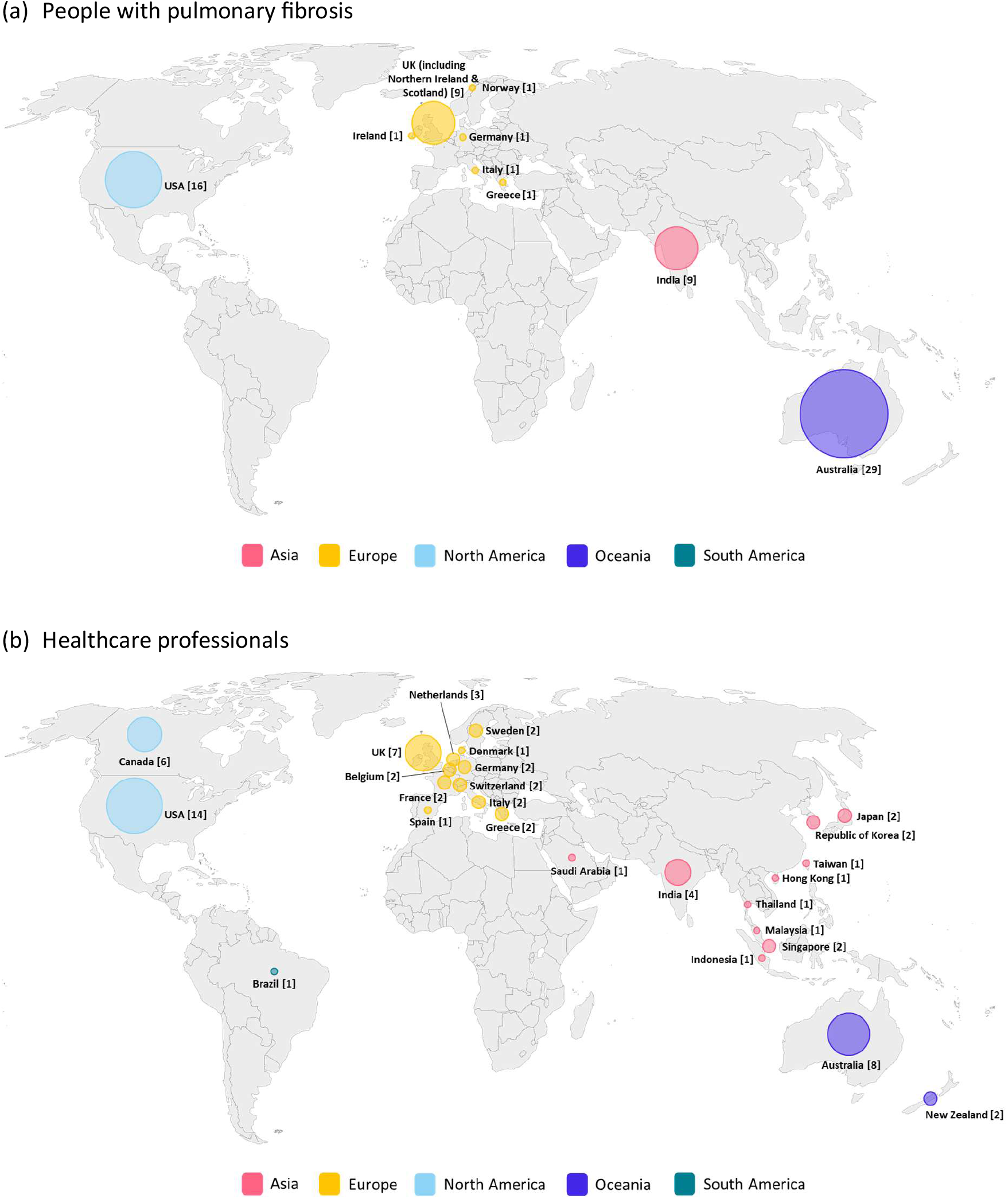
Represented countries for (a) people with pulmonary fibrosis and (b) healthcare professionals in Part A survey Abbreviations: UK, United Kingdom; USA, United States of America

### Patient experience and current clinical practice

Forty-nine percent of people with PF reported having discussions about nocturnal hypoxaemia, which were initiated primarily by the lung specialist (48%), patients themselves (33%), or jointly (3%) (**Figure 2** and **Table S1**). Previous investigations into nocturnal hypoxaemia occurred in 51% of patients, which included polysomnography (69%), overnight oximetry (49%), and wearables (9%), with some reporting discomfort or difficulty sleeping with polysomnography. The majority of patients (75%) who had not previously completed sleep-related investigation for nocturnal hypoxaemia indicated it should be part of their disease assessment for pulmonary fibrosis.

**Figure 2.**
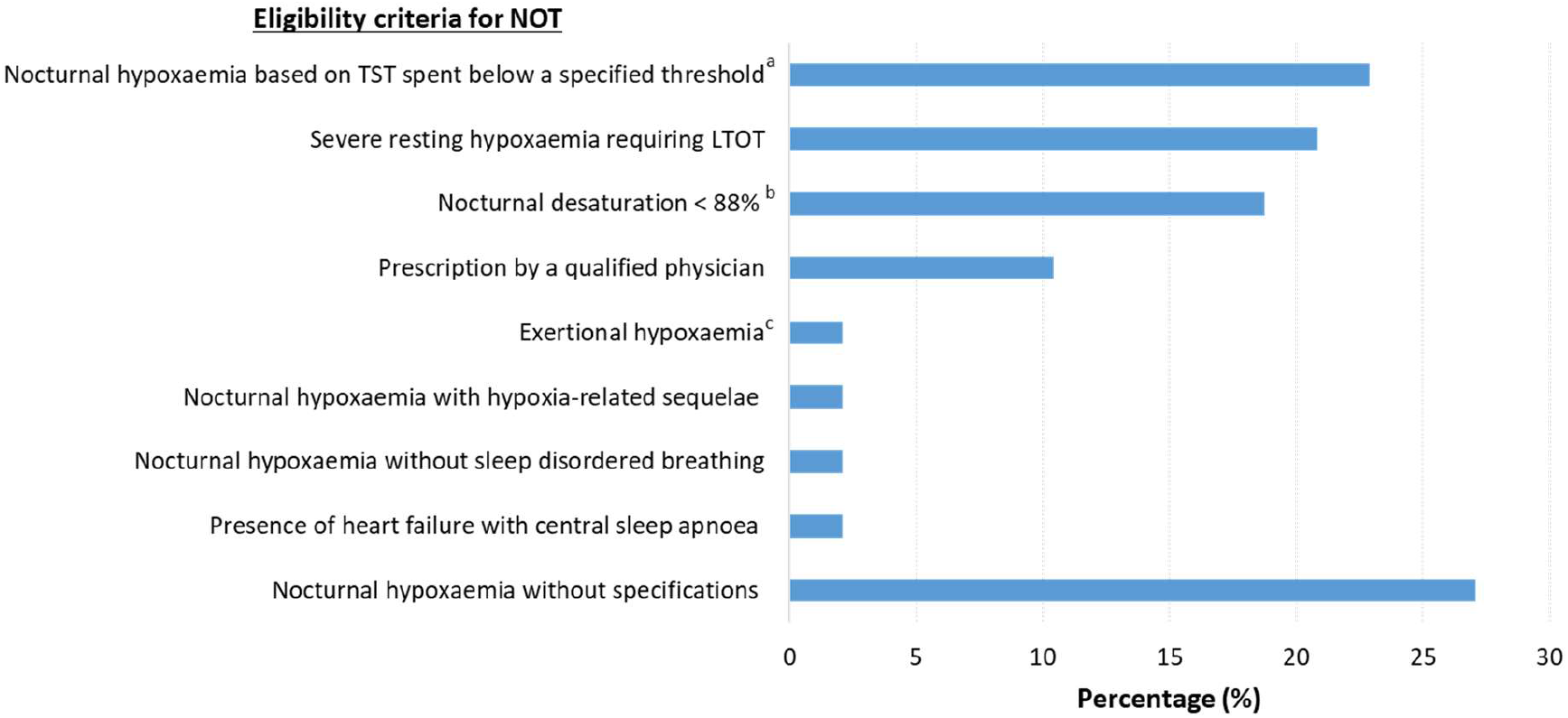
Eligibility criteria for nocturnal oxygen therapy Abbreviations: LTOT, long-term oxygen therapy; TST, total sleep time ^*^ Based on responses from 48 healthcare professionals who had prescribed nocturnal oxygen therapy for people with pulmonary fibrosis ^a^ Different thresholds for the degree (80-88%) and duration (20-33%) of nocturnal desaturation were described, with some requiring additional criteria such as presence of hypoxia-related sequelae or correction with supplemental oxygen therapy ^b^ Different thresholds of nocturnal desaturation were described, with some requiring the presence of hypoxia-related sequelae ^C^ One reported only eligible in the context of being evaluated or waitlisted for lung transplant

Of the patients who were current or previous users of NOT, the therapy was funded by the government (45%), patients themselves (21%), private health insurance (14%), hospital (3%), or with joint payments by the government and private health insurance or patients (17%). Most patients (76%) reported regular daily use of NOT. Some patients tolerated NOT well without side effects and reported improved sleep quality and reduced fatigue or headache. Reported side effects of NOT included noise of the oxygen concentrator, nasal irritation, and fall risk with nasal cannula. For patients who had never used NOT, 50% would use NOT if required and 26% were interested to learn more about NOT. In addition to oxygen prescription, patients expressed the need to receive information on the indication, benefits, and side effects of NOT, as well as tolerability and practical tips for its use, including ways to improve nasal comfort and avoid nasal cannula dislodgement, as important aspects to know before treatment commencement.

Among HCPs, 64% and 48% performed assessments for sleep and nocturnal hypoxaemia routinely or as clinically indicated, respectively (**Table S2**). Both polysomnography and overnight oximetry were used for routine assessment of nocturnal hypoxaemia, with 59% being conducted at baseline. The main reasons for not routinely assessing nocturnal hypoxaemia in PF were limited resources available for polysomnography and/or overnight oximetry (40%) and the lack of high-level evidence for treating nocturnal hypoxaemia (21%). Common triggers for assessing nocturnal hypoxaemia were suspected or confirmed OSA or other sleep disturbance (74%), daytime resting or exertional hypoxaemia (55%), and the presence of pulmonary hypertension (30%). Nevertheless, the vast majority of HCPs had access to polysomnography (90%) or overnight oximetry (89%).

Of the HCPs who were involved in patient care, 89% reported that they initiated discussions on nocturnal hypoxaemia during clinical consultations most of the time or more often than their patients. Eighty-two percent of HCPs indicated that assessment of nocturnal hypoxaemia as very or somewhat important in people with PF due to the association of nocturnal hypoxaemia with pulmonary hypertension and poor survival. Most HCPs provided continuous positive airway pressure (CPAP) for OSA in people with pulmonary fibrosis, with NOT being considered as a sole or additional intervention in some (23%). Over 70% of HCPs had access to funded NOT for people with pulmonary fibrosis, although the eligibility criteria for NOT substantially varied (**Figure 2**). Treating nocturnal hypoxaemia was considered to be at somewhat important for improving patient outcomes in the majority of HCPs (64%), with others expressing the need for more research (13%), lack of benefits in the absence of daytime hypoxaemia (13%), and uncertainties (10%).

### Research priorities on nocturnal hypoxaemia in PF

Top patients’ and healthcare professionals’ research priorities on nocturnal hypoxaemia in PF are presented in **Table 3**, with detailed results in **Tables S3** and **S4**. Among people with pulmonary fibrosis, top research priorities on nocturnal hypoxaemia identified were the effects of its treatment on symptom burden, tolerability of NOT for patients and their partners, developing a user-friendly oxygen therapy device, side effects of using NOT, patients’ understanding of the significance of nocturnal hypoxaemia, and diagnostic approaches for assessing sleep and nocturnal hypoxaemia.

**Table 3.** Top patients’ and healthcare professionals’ research priorities on nocturnal hypoxaemia in pulmonary fibrosis.

| People with pulmonary fibrosis | Healthcare professionals |
| --- | --- |
| Effects of treating nocturnal hypoxaemia on symptom burden of pulmonary fibrosis | Effects of treating nocturnal hypoxaemia on survival |
| Tolerability of nocturnal oxygen therapy for patients and their partners | Effects of treating nocturnal hypoxaemia on health-related quality of life |
| Developing a user-friendly oxygen therapy device | Effects of treating nocturnal hypoxaemia on pulmonary hypertension |
| Side effects of using nocturnal oxygen therapy | Predictors of nocturnal hypoxaemia |
| Patients' understandings of the significance of nocturnal hypoxaemia | Thresholds to initiate treatment for nocturnal hypoxaemia |
| Diagnostic approaches for assessing sleep and nocturnal hypoxaemia | Diagnostic approaches for assessing sleep and nocturnal hypoxaemia |
| Effects of treating nocturnal hypoxaemia on cognitive performance* | Effects of treating nocturnal hypoxaemia on cognitive performance* |

\* This was added as a research priority by both people with pulmonary fibrosis and healthcare professionals during online focus groups.

Most research priorities on nocturnal hypoxaemia in PF raised by HCPs were related to its treatment effects, including survival, HRQoL, and development and/or progression of pulmonary hypertension. In addition to the shared priority of diagnostic approaches for sleep and nocturnal hypoxaemia, HCPs also identified establishing predictors and thresholds to initiate treatment for nocturnal hypoxaemia in PF as research priorities. Other research areas that were raised by both people with PF and HCPs included comparison of effects of different treatment modalities for nocturnal hypoxaemia, such as NOT, continuous positive airway pressure (CPAP), and non-invasive ventilators, delivery of patient education and support, utility of wearables and questionnaires for screening and assessment, costs and accessibility of NOT, and relationships between nocturnal hypoxaemia and sleep disturbance with health outcomes in pulmonary fibrosis.

During the focus group discussions, top research priorities raised by people with PF and HCPs were endorsed. Treatment effects of nocturnal hypoxaemia on symptom burden including HRQoL, as well as usability and tolerability of NOT, were emphasised by both groups. Key aspects of symptom burden were discussed, with sleep quality and daytime physical and cognitive function being prioritised. The importance of assessing cognitive performance was stressed by people with PF due to concerns about sleep disturbance and nocturnal hypoxaemia on memory, attention span, and problem solving that could impact disease coping for pulmonary fibrosis, which was endorsed by HCPs and added as a top research priority. The need to study the differences in health outcomes and treatment effects for nocturnal hypoxaemia with and without co-existing OSA or other sleep disordered breathing in PF was raised by HCPs.

### Outcomes measures for evaluation of NOT in PF

For people with pulmonary fibrosis, the median score for all 16 listed outcome measures was 4 “very important”, with forced vital capacity (FVC), PF-specific and sleep-specific HRQoL, apnoea-hypopnoea index (AHI), and oxygenation status during sleep being rated such by ≥ 80% of respondents (**Table 4**). On the other hand, the median scores were 4 “very important” for 11 out of 16 listed outcome measures among HCPs, with the lower-scoring ones being FVC, diffusion capacity of carbon monoxide, depression, AHI, and sleep efficiency (**Table 4**). Only two outcome measures, survival and PF-specific HRQoL, were rated as “very important” in ≥ 80% of HCPs. Additional outcomes were nominated by people with PF and HCPs (**Table 5**).

**Table 4.** Assessment of proposed outcomes measures for evaluation of nocturnal oxygen therapy in pulmonary fibrosis.

| Outcome measures | People with pulmonary fibrosis<br>(n = 68) |  | Healthcare professionals<br>(n = 73) |  |
| --- | --- | --- | --- | --- |
|  | Median<br>(IQR) | Very important<br>(%) | Median<br>(IQR) | Very important<br>(%) |
| **ILD-related outcomes** |  |  |  |  |
| Survival | 4 (1) | 65 | 4 (0) | 89 |
| FVC | 4 (0) | 80 | 3 (1) | 19 |
| DLCO | 4 (1) | 71 | 3 (2) | 26 |
| Disease progression | 4 (1) | 75 | 4 (1) | 52 |
| Development/progression of PH | 4 (0.75) | 75 | 4 (0) | 79 |
| Development of resting hypoxaemia | 4 (0) | 75 | 4 (1) | 52 |
| **Patient-reported outcomes** |  |  |  |  |
| PF-specific HRQoL | 4 (0) | 80 | 4 (0) | 84 |
| Sleep-specific HRQoL | 4 (1) | 83 | 4 (0.5) | 75 |
| Sleep quality | 4 (1) | 72 | 4 (1) | 67 |
| Daytime breathlessness | 4 (0.75) | 75 | 4 (1) | 52 |
| Daytime fatigue | 4 (1) | 73 | 4 (1) | 66 |
| Anxiety | 4 (2) | 56 | 4 (1) | 47 |
| Depression | 4 (2) | 58 | 3 (1) | 47 |
| **Objective sleep parameters** |  |  |  |  |
| AHI | 4 (0) | 83 | 3 (1) | 34 |
| Oxygenation status during sleep | 4 (0) | 87 | 4 (1) | 71 |
| Sleep efficiency | 4 (1) | 72 | 3 (1) | 41 |

Abbreviations: AHI, apnoea-hypopnoea index; DLCO, diffusing capacity for carbon monoxide; FVC, forced vital capacity; HRQoL, health-related quality of life; PF, pulmonary fibrosis; PH, pulmonary hypertension

**Table 5.** Additional outcome measures for evaluation of nocturnal oxygen therapy in pulmonary fibrosis nominated by people with pulmonary fibrosis and healthcare professionals.

| Outcome measures | People with pulmonary fibrosis | Healthcare professionals |
| --- | --- | --- |
| PF-related outcomes | • Hospitalisation<br>• Acute exacerbation | • Hospitalisation<br>• Acute exacerbation<br>• Functional exercise capacity |
| Patient-reported outcomes | • Cognition<br>• Impact on carers, partners, and/or family | • Social impact |
| Safety | • Side effects | • Side effects |
| Other | • Other organ/system outcomes (including cognitive performance) | • Cardiovascular outcomes<br>• Cost-effectiveness |

Abbreviations: PF, pulmonary fibrosis;

During the online focus groups, discrepancies in the top-rated listed outcome measures by people with PF and HCPs were discussed. It was acknowledged that objective measures of FVC and AHI were the primary focus for evaluation of PF and OSA severities discussed during clinical consultations, respectively, with the former being a measure for determining disease progression PF as well. Consensus was achieved across both groups that HRQoL was the key outcome measure of interest amongst the top-rated ones. Of the additional nominated outcomes, cognitive performance was selected for inclusion as an important measure.

## Discussion

This study provides in-depth insights into stakeholders’ experiences and perceptions, as well as research priorities and key outcome measures, to inform future research on sleep assessment and treatment of nocturnal hypoxaemia in pulmonary fibrosis. While there were wide variations in current clinical care for the diagnosis and management of sleep disturbance and nocturnal hypoxaemia in pulmonary fibrosis, significant proportions of patients and HCPs expressed their importance as part of the disease management in pulmonary fibrosis. Both people with PF and HCPs collectively identified treatment effects of nocturnal hypoxaemia on symptom burden and HRQoL in PF and the usability and tolerability of NOT as top research priorities, with HRQoL being the key outcome measure of interest in the evaluation of NOT. Relationships between cognitive performance with sleep disturbance and nocturnal hypoxaemia were raised by patients and endorsed by HCPs as a top research area in pulmonary fibrosis.

Despite advancements in disease-targeted therapies with the availability of antifibrotic medications since the mid-2010s, people with PF continue to experience significant morbidity and mortality.^8^ These ongoing unmet patients’ needs emphasise the importance of comprehensive care, given that disease manifestations such as hypoxaemia and comorbidities can impact symptom burden and health outcomes in people with pulmonary fibrosis.^9^ Sleep is an essential indispensable physiological requirement for daily restoration and homeostasis of body systems, with nocturnal hypoxaemia and OSA commonly affecting people with pulmonary fibrosis.^1,3^ We found that clinical discussion and investigation of sleep disturbance and nocturnal hypoxaemia occurred in over 50% of people with pulmonary fibrosis, although the tests and timing of assessment varied substantially. Infrastructure availability and accessibility to required tests and interventions are known barriers to optimal care for pulmonary fibrosis.^10^ It is noteworthy that the prescribing and funding criteria and funding mechanisms for NOT differ across different regions, with over 27% having no access requiring self- funding by patients. Although the vast majority of HCPs had access to polysomnography or overnight oximetry for sleep assessment, the available resources could be restricted.

As raised by HCPs, there are currently limited high-level data to guide evidence-based clinical practice in sleep assessment and treatment of NOT in pulmonary fibrosis, despite the perceived significant relationships and potential therapeutic benefits. This was a key factor of inconsistent clinical practice and likely also contributes to differences in clinical guideline recommendations and policies on NOT across jurisdictions. Research into the effects and delivery of supplemental oxygen therapy has consistently been identified as stakeholders’ priorities in pulmonary fibrosis.^11,12^

Furthermore, access to supplemental oxygen therapy is a key area of inequality and unmet need in pulmonary fibrosis.^13^ However, it is important to acknowledge the potential patient and carer burden associated with oxygen use.^14-16^ Nocturnal interventions for OSA, such as CPAP, are reported to be challenging in people with PF due to altered breathing patterns, cough symptoms, and insomnia secondary to drug therapy or comorbidities.^17^ While NOT appears to be less intrusive than CPAP, its use is associated with fall risk, concerns of nasal cannula dislodgement, and sleep disturbance from discomfort or noise generated by the oxygen concentrators that may also affect sleep partners.^18^ Thorough evaluation of both potential treatment effects and associated negative impact of NOT in people with PF is required to inform optimal clinical care and appropriate patient education and support.

In the survey, there were some discrepancies in the top research priorities on nocturnal hypoxaemia and NOT, as well as top-rated outcome measures for NOT, between people with PF and HCPs, which may be related to differences in the perceptions and lived experiences of people with PF compared to HCPs. The potential impact of nocturnal hypoxaemia and NOT on pulmonary hypertension was emphasised by HCPs, given the physiological plausibility and prognostic significance of pulmonary hypertension in pulmonary fibrosis.^1,19^ Following online focus groups, symptom burden and HRQoL were identified as the top research priority on nocturnal hypoxaemia and NOT, with both groups highlighting how patients feel being one of the critical determinants in clinical care. The effects of sleep disturbance and nocturnal hypoxaemia on cognitive performance were raised by people with PF as a top research priority. There is recent recognition of increased cognitive impairment including deficits in executive functioning and processing speed in people with ILD in case-control studies,^20^ which warrants targeted research given the importance of cognitive performance in self-management and daily tasks.

While participation in this study was limited to English-speaking stakeholders, we had global engagement for both people with PF and HCPs of different backgrounds and regions. The HCP survey participation rate was high at 68% (n=73/108), although the patient survey participation rate could not be determined due to the recruitment strategy. People with PF and HCPs who completed this study were likely to have greater interest in sleep health and NOT, compared to those who did not participate. Nevertheless, a range of experiences and perceptions were identified in our study, indicating its representativeness and relevance for wide application.

## CONCLUSION

Our study highlights the varying clinical care and challenges in sleep assessment and treatment of nocturnal hypoxaemia in people with PF, despite the broad acceptance of its significance and potential impact on the disease course and health outcomes of PF. Top research priorities on nocturnal hypoxaemia and NOT, as well as key outcome measures for evaluation of NOT, established by international stakeholders provide guidance for future research efforts. Interdisciplinary collaboration is crucial to optimise research efforts to address and understand the complexity of the mechanisms underlying the association and interactions between PF, sleep disturbance, and nocturnal hypoxaemia, as well as the effects of NOT and other positive airway pressure interventions.

## Data Availability

All data produced in the present study are available upon reasonable request to the authors

## ACKNOWLEDGEMENT

This study was funded by Medical Research Future Fund (ID: 2030893). YHK receives fellowship support from the National Health and Medical Research Council Investigator Grant (ID: 2008255). The authors thank for Lung Foundation Australia, European Pulmonary Fibrosis Federation, and PF Warriors for recruitment assistance, as well as all participants.

