## Supplementary material for "International stakeholder engagement on clinical care and research for nocturnal hypoxaemia in pulmonary fibrosis": Table S1, Table S2, Tables S3 and S4

**Table S1**. Patients' experiences and views regarding sleep assessment and treatment for nocturnal hypoxaemia in pulmonary fibrosis

| Questions | N | Responses |
|---|---|---|
| Have you and your treating doctor(s) ever discussed assessing and/or treating you for low oxygen level during sleep? | 68 | • Yes = 33<br>• No = 33<br>• Unsure = 2 |
| Who raised the discussion about assessing and treating for low oxygen level during sleep in clinic consultations? (Choose all that apply) | 33 | • Lung specialist = 16<br>• Patient = 11<br>• Both patient and lung specialist = 1<br>• Other = 5<br>− Cardiologist: 2<br>− Primary care physician: 1<br>− Otolaryngologist: 1<br>− Sleep physician: 1 |
| Have you ever had testing for low oxygen level during sleep or for use of oxygen therapy during sleep (e.g. staying overnight in hospital or sleep laboratory, or taking a device or oximeter home for wearing overnight)? | 68 | • Yes = 35<br>• No = 33 |
| Which test(s) did you have? (Choose all that apply) | 35 | • PSG = 24<br>• Overnight oximetry = 17<br>• Wearables = 3<br>• Not stated = 1 |
| Can you tell us about your experience with doing the test(s)? (For patients who have had sleep testing) | 34 | **Overnight oximetry n = 16**<br>• Easy = 4<br>• Informative = 4<br>• Remote set-up = 2<br>• A little annoying = 1<br><br>**PSG, n = 23**<br>• Uncomfortable/difficulty sleeping = 10<br>• Informative = 6<br>• Easy = 4<br>• Manageable = 3<br>• Remote set-up = 2<br><br>**Wearables n = 4**<br>• Easy = 2<br>• Informative = 1 |
| What do you think about having tests for low oxygen level during sleep as part of your assessment for pulmonary fibrosis? (For patients who haven't had sleep testing) | 32 | • Yes = 21<br>• Yes, although need to know more = 3<br>• No = 5<br>• If recommended by my doctor = 1<br>• If it's overnight oximetry = 1<br>• Unsure = 1 |
| What would you want to know before starting oxygen therapy during sleep? | 63 | • Treatment benefits (including consequences if untreated) = 15<br>• Tolerability of NOT = 11<br>• Treatment side effects = 11<br>• How to use NOT = 11<br>• Indication for NOT = 9 |

| | | |
|---|---|---|
| | | • Practical tips for using NOT = 7<br>• How to use with CPAP = 5<br>• Monitoring required = 3<br>• Financial cost = 2<br>• Impact on bed partner = 2<br>• Patient education provision = 2<br>• Risks of hypercapnoea = 2<br>• Accessibility = 1<br>• Shared experience from current user = 1 |
| **For patients who were current and previous users of nocturnal oxygen therapy** | | |
| Who pays/paid for nocturnal oxygen therapy? | 29 | • Government: 13<br>• Self = 6<br>• Joint payment = 5 (Government + private health insurance or self)<br>• Private health insurance: 4<br>• Hospital = 1 |
| Can you tell us about your experience with using oxygen therapy during sleep? (For examples, how often do you use it, how many hours per night, any side effects, etc) | 29 | • Using regularly every night and/or during sleep = 22<br>• Intermittent use = 2<br>• No side effects = 8<br>• Improved sleep quality = 5<br>• Reduced fatigue = 2<br>• Less headache = 2<br>• Side effects = 3<br>– Noise from the concentrator<br>– Nasal dryness and irritation<br>– Falls risk with oxygen tubing |
| **For patients who had never used nocturnal oxygen therapy** | | |
| What do you think about using oxygen therapy during sleep? | 38 | • Would use if needed or sleep better: 19<br>• Interested to learn more = 10<br>• Unsure = 3<br>• Unkeen = 3<br>• Would use if not affecting partner = 1<br>• Would use for part of the night = 1<br>• No = 1 |

Abbreviations: CPAP, continuous positive airway pressure; NOT, nocturnal oxygen therapy; PSG, polysomnography

**Table S2**. Healthcare professionals' experiences and views regarding sleep assessment and treatment for nocturnal hypoxaemia in pulmonary fibrosis

| Questions | N | Responses |
|---|---|---|
| Do you routinely assess sleep in people with pulmonary fibrosis? | 73 | • Yes = 27<br>• No = 26<br>• Only indicated = 20<br>− Suspected OSA<br>− Daytime hypoxaemia<br>− Exertional hypoxaemia |
| Do you routinely assess for nocturnal hypoxaemia in people with pulmonary fibrosis? | 73 | • No = 38<br>• Only indicated = 18<br>− Suspected OSA<br>− Exertional hypoxaemia<br>− Progressive ILD<br>− Disproportionate pulmonary hypertension<br>− Hypercapnoea<br>− Daytime hypoxaemia<br>• Yes = 17 |
| For routine assessment of nocturnal hypoxaemia, which test(s) do you order? (Choose all that apply) | 17 | • Overnight oximetry = 15<br>• PSG = 9 |
| For routine assessment of nocturnal hypoxaemia, how often do you perform the test(s)? | 17 | • Other = 13<br>− As indicated (e.g. oxygen therapy assessment) = 7<br>− Baseline and as indicated = 3<br>− Baseline and at intervals = 2 (vary for different patients)<br>− Baseline and when considering long-term oxygen therapy = 1<br>• Baseline = 2<br>• Baseline and annually = 2 |
| Why do you not routinely assess for nocturnal hypoxaemia in people with pulmonary fibrosis? | 52 | • Limited resource available for routine testing (e.g. need hospital admission, refer to a different facility/services) = 21<br>• No/limited evidence on benefits for correcting nocturnal hypoxaemia = 11<br>• Only indicated for patients with suspected nocturnal hypoxaemia = 9<br>• Not part of standard ILD care = 6<br>• Costs associated with tests for sleep assessments = 3<br>• Limited access to NOT = 3<br>• Perceived patient reluctance to testing or NOT = 3<br>• Sleep evaluation performed by other physicians = 3<br>• Only for long-term oxygen therapy prescription = 1 |
| What will trigger your assessment for nocturnal hypoxaemia in people with pulmonary fibrosis? | 53 | • Suspected or presence of OSA or other sleep disturbance = 39<br>• Resting and/or exertional hypoxaemia = 29<br>• Presence of pulmonary hypertension = 16<br>• Evidence supporting benefits for treating nocturnal hypoxaemia = 6<br>• At-risk of pulmonary hypertension = 5<br>• Excessive daytime symptoms (dyspnoea, fatigue, reduced exercise capacity) = 3<br>• Assessment for oxygen therapy = 4<br>• Severe ILD = 3 |

| | | |
|---|---|---|
| | | • Presence of hypoxia-related sequelae = 3<br>• Progressive ILD = 2<br>• Self-monitoring data = 1 |
| Do you have access to tests for assessing nocturnal hypoxaemia? | 72 | • Polysomnography: Yes = 65; No = 7<br>• Overnight oximetry: Yes = 64; No = 8<br>• Other tests:<br>− Transcutaneous carbon dioxide monitoring = 3<br>− 24-hour home oximetry = 1<br>− WatchPat one = 1<br>− Actigraphy = 1 |
| Who usually raises the idea of assessing for nocturnal hypoxaemia when discussed in clinic consultations? | 66 | • I raise the idea of the test most or all of the time = 38<br>• I raise the idea of the test more often than the patient = 21<br>• The patient raises the idea of the test more often than me = 6<br>• The patient raises the idea of the test most or all of the time = 1 |
| In your opinion, how important is assessing nocturnal hypoxaemia in people with pulmonary fibrosis? | 73 | • Very important = 29<br>• Somewhat important = 31<br>• Neutral = 4<br>• Somewhat unimportant = 7<br>• Not important at all = 2 |
| Provide comments on your perceived importance on assessing nocturnal hypoxaemia in people with pulmonary fibrosis. | 64 | **Very or Somewhat important**<br>• Association with pulmonary hypertension = 21<br>• Prognostic value for survival = 19<br>• Treatment benefits for health-related quality of life = 12<br>• Assessment for oxygen therapy = 11<br>• Association with OSA or other sleep disordered breathing = 10<br>• Association with cardiovascular events = 6<br>• Treatment benefits for dyspnoea and exercise capacity = 6<br>• Available interventions for nocturnal hypoxaemia = 3<br>• Treatment benefits for fatigue = 2<br>• Extrapolated from COPD and heart failure patients = 1<br>• Treatment benefits for cognition = 1<br>• Cannot be identified by clinical assessment = 1<br>• Treatment benefits for sleep quality = 1<br><br>**Neutral**<br>• May be in selected populations = 2<br>• Lack of evidence although suspected scientific importance = 1<br>• No evidence in ILD and no benefits seen in COPD = 1<br><br>**Somewhat unimportant or Not important at all**<br>• Lack of evidence for treatment or prognostication = 6<br>• Unclear benefits = 4 |
| How do you treat sleep apnoea in people with pulmonary fibrosis? | 66 | • CPAP or APAP = 28<br>• Treatment as per standard care for OSA = 15<br>• Treatment provided by another specialist = 8<br>• CPAP +/- oxygen = 7<br>• CPAP or NIV = 5<br>• CPAP or oxygen = 3<br>• CPAP or NIV +/- oxygen = 2 |

| | | |
|---|---|---|
| | | • NOT = 2<br>• NIV = 2<br>• Under-resourced to treat = 2<br>• Additional monitoring of echocardiogram = 1<br>• NIV +/- oxygen = 1 |
| Do you have access to funded nocturnal oxygen therapy in people with pulmonary fibrosis? | 72 | • Yes = 52<br>• No = 20 |
| What are the requirements for funded nocturnal oxygen therapy in people with pulmonary fibrosis? | 48 | • Nocturnal hypoxaemia without specifications = 13<br>• Nocturnal hypoxaemia meeting a threshold based on total sleep time spent at 88% or less with and without additional qualifiers = 11<br> − TST ≤ 88% of 30% or 1/3 of night = 8<br> − TST ≤ 88% of 20% = 1<br> − TST ≤ 88% of 30% and correction with supplemental oxygen = 1<br> − TST ≤ 88% of 30% or 1/3 of night with hypoxia-related sequelae = 1<br> − TST ≤ 80% of 10% = 1<br>• Criteria for long-term oxygen therapy = 10<br>• Nocturnal desaturation < 88% with and without additional qualifiers = 9<br> − Desaturation duration > 5 mins = 4<br> − Nocturnal desaturation < 80%<br> − Presence of hypoxia-related sequelae = 1<br> − Sustained desaturation, duration note specified = 1<br>• Prescription from a qualified physician = 5<br>• Exertional hypoxaemia with and without additional qualifiers = 2<br> − Exertional hypoxaemia and being evaluated or waitlisted for lung transplant = 1<br>• Nocturnal hypoxaemia with hypoxia-related sequelae = 1<br>• Nocturnal hypoxaemia without sleep disordered breathing = 1<br>• Presence of heart failure with central sleep apnoea = 1<br>• Uncertain = 1 |
| In your opinion, how useful is treating nocturnal hypoxaemia in people with pulmonary fibrosis? | 69 | • Very important = 18<br>• important = 16<br>• Somewhat important = 10<br>• Need more data = 9<br>• Not helpful in the absence of daytime hypoxaemia = 9<br>• Unclear = 7<br><br>Potential beneficial effects described include:<br>• Pulmonary hypertension = 14<br>• Health-related quality of life = 11<br>• Cardiovascular events (including heart failure) = 6<br>• Survival = 6<br>• Overall morbidity = 5<br>• Sleep quality = 5<br>• Hospitalisation = 1<br>• Physical activities = 1<br>• Polycythaemia = 1 |

Abbreviations: APAP, auto-adjustable positive airway pressure; COPD, chronic obstructive pulmonary disease; ILD, interstitial lung disease; NIV, non-invasive ventilation; NOT, nocturnal oxygen therapy; OSA, obstructive sleep apnoea; PSG, polysomnography; TST, total sleep time

**Table S3**. Patients' research priorities on nocturnal hypoxemia in pulmonary fibrosis (n = 68)

| Topics | N (%) |
|---|---|
| **Effects of treating nocturnal hypoxaemia** | |
| • Effects of treating nocturnal hypoxaemia on symptom burden of pulmonary fibrosis | 23 |
| • Effects of treating nocturnal hypoxaemia on cardiovascular outcomes | 3 |
| • Effects of treating nocturnal hypoxaemia on quality of life | 1 |
| • Effects of treating nocturnal hypoxaemia on ILD disease progression | 2 |
| • Effects of treating nocturnal hypoxaemia on OSA | 1 |
| • Effects of treating nocturnal hypoxaemia on survival | 1 |
| • Effects of treating nocturnal hypoxaemia in different ILD severity | 1 |
| • Effects of treating nocturnal hypoxaemia for different altitude levels | 1 |
| **Feasibility of delivering nocturnal oxygen therapy** | |
| • Tolerability of nocturnal oxygen therapy for patients and their partners | 21 |
| • Developing a user-friendly oxygen therapy device | 20 |
| • Side effects of using nocturnal oxygen therapy | 16 |
| • Monitoring of nocturnal oxygen therapy use | 8 |
| • Accessibility to nocturnal oxygen therapy | 8 |
| • Effective titration of nocturnal oxygen therapy | 7 |
| • Financial cost of nocturnal oxygen therapy | 7 |
| • Education and support for nocturnal oxygen therapy | 3 |
| **Assessment for sleep disturbance and nocturnal hypoxaemia** | |
| • Diagnostic approaches for assessing sleep and nocturnal hypoxaemia | 9 |
| • Utility of wearables for screening and assessing sleep and nocturnal hypoxaemia | 6 |
| • Value of the involvement of sleep specialists in ILD care | 2 |
| **Epidemiology of sleep disturbance and nocturnal hypoxaemia** | |
| • Predictors of nocturnal hypoxaemia | 5 |
| • Prevalence of nocturnal hypoxaemia | 4 |
| • Factors contributing to the development of nocturnal hypoxaemia | 3 |
| • Prevalence of OSA | 2 |
| **Awareness of nocturnal hypoxaemia** | |
| • Patients' understandings on the significance of nocturnal hypoxaemia | 10 |
| • Clinicians' understandings on the significance of nocturnal hypoxaemia | 5 |
| **Treatment options for nocturnal hypoxaemia** | |
| • Comparison of nocturnal oxygen therapy and CPAP | 7 |
| **Effects of sleep disturbance and nocturnal hypoxaemia** | |
| • Effects of nocturnal hypoxaemia on health outcomes | 6 |
| • Effects of OSA on health outcomes | 2 |

Abbreviations: CPAP, continuous positive airway pressure; ILD, interstitial lung disease; obstructive sleep apnoea

**Table S4**. Healthcare professionals' research priorities on nocturnal hypoxemia in pulmonary fibrosis (n = 73)

| Topics | N (%) |
|---|---|
| **Effects of treating nocturnal hypoxaemia** | |
| • Effects of treating nocturnal hypoxaemia on survival | 20 |
| • Effects of treating nocturnal hypoxaemia on health-related quality of life | 14 |
| • Effects of treating nocturnal hypoxaemia on pulmonary hypertension | 14 |
| • Effects of treating nocturnal hypoxaemia on symptom burden of pulmonary fibrosis | 11 |
| • Effects of treating nocturnal hypoxaemia on ILD disease progression | 4 |
| • Effects of treating nocturnal hypoxaemia on cardiovascular outcomes | 4 |
| • Effects of treating nocturnal hypoxaemia on daytime oxygenation status | 4 |
| **Feasibility of delivering nocturnal oxygen therapy** | |
| • Thresholds to initiate treatment for nocturnal hypoxaemia | 14 |
| • Side effects of using nocturnal oxygen therapy | 3 |
| • Tolerability of nocturnal oxygen therapy for patients | 3 |
| • Effective titration of nocturnal oxygen therapy | 2 |
| • Cost-effectiveness of nocturnal oxygen therapy | 2 |
| • Accessibility to nocturnal oxygen therapy | 2 |
| • Education and support for nocturnal oxygen therapy | 2 |
| **Epidemiology of sleep disturbance and nocturnal hypoxaemia** | |
| • Predictors of nocturnal hypoxaemia | 14 |
| • Prevalence of nocturnal hypoxaemia | 4 |
| • Prevalence of OSA | 2 |
| **Effects of sleep disturbance and nocturnal hypoxaemia** | |
| • Effects of nocturnal hypoxaemia without OSA on survival | 11 |
| • Effects of nocturnal hypoxaemia without OSA on health-related quality of life | 5 |
| • Effects of OSA on health outcomes | 1 |
| **Assessment for sleep disturbance and nocturnal hypoxaemia** | |
| • Diagnostic approaches for assessing sleep and nocturnal hypoxaemia | 14 |
| • Utility of wearables for screening and assessing sleep and nocturnal hypoxaemia | 6 |
| • Utility of questionnaires for screening and assessing sleep and nocturnal hypoxaemia | 2 |
| **Treatment options for nocturnal hypoxaemia** | |
| • Comparison of nocturnal oxygen therapy and CPAP | 2 |
| • Comparison of nocturnal oxygen therapy and non-invasive ventilator | 2 |
| • Comparison of nocturnal oxygen therapy and high-flow oxygen therapy | 1 |

Abbreviations: CPAP, continuous positive airway pressure; ILD, interstitial lung disease; obstructive sleep apnoea
